# The Congenital Heart Initiative: Results from a Patient-Centered Registry for Adults with Congenital Heart Disease

**DOI:** 10.1101/2024.05.16.24307510

**Authors:** Scott Leezer, Mindi Messmer, Anushree Agarwal, Ruth Phillippi, Jamie L. Jackson, Mark Roeder, Aliza Marlin, Noah Peyser, Mark J. Pletcher, Richard Krasuski, Matthew Lewis, Leigh Reardon, Arwa Saidi, Ronald Kanter, Satinder Sandhu, Thomas Young, Roni Jacobsen, Emily Ruckdeschel, Adam Lubert, Simran Singh, Ali Zaidi, Dan H. Halpern, Anita Mathews, Rittal Mehta, Thomas Carton, Anitha S. John

**Affiliations:** CURA Strategies, Arlington, VA; Children’s National Hospital, Washington, DC; University of California San Francisco, San Francisco, CA; Nationwide Children’s Hospital, Columbus, OH; Adult Congenital Heart Association, Philadelphia, PA; Duke University Health System, Durham, NC; Columbia University Irving Medical Center, New York, NY; University of California Los Angeles, Los Angeles, CA; University of Florida, Gainesville, FL; Nicklaus Children’s Hospital, Miami, FL; University of Miami, Miami, FL; Ochsner Health, New Orleans, LA; Children’s Hospital of Colorado, Denver, CO; Children’s Hospital of Philadelphia, Philadelphia, PA; Cincinnati Children’s Hospital, Cincinnati, OH; Weill Cornell Medicine, New York, NY; Mt. Sinai Hospital, New York, NY; NYU Langone Health, New York, NY; Louisiana Public Health Institute, New Orleans, LA

**Keywords:** Congenital Heart Disease, Digital Health, Registry, Patient Reported Outcomes

## Abstract

**Background:** In the United States, there are over 1.5 million adults living with congenital heart disease (CHD). The Congenital Heart Initiative (CHI) is a digital, online patient empowered registry that was created to advance multicenter research and improve clinical care by gathering patient-reported outcomes (PROs) in adults with CHD.

**Methods:** After a two year design process, the CHI was created and launched nationally on December 7, 2020 using a human centered design approach. Demographics and validated survey tools on quality of life, mental health, physical activity and health care utilization were collected at baseline and every 4 months. Data were collected virtually and stored on Health Insurance Portability and Accountability Act (HIPAA)–compliant cloud-based servers with restricted access.

**Results:** By December 31, 2023, the CHI had enrolled 4558 participants (56% female) with an average age of 39 years ± 14, representing all 50 states. Approximately 88% of participants have completed at least one e-Visit as of December 31, 2023. The most prevalent CHD anatomy included tetralogy of Fallot (883, 22%), transposition of great arteries (452, 11%), and coarctation of the aorta (429, 11%). Approximately 88% of participants reported at least one co-morbidity, with arrhythmia (1310, 29%) and mood disorder (1339, 29%) as the most common cardiac and non-cardiac co-morbidity, respectively. Among female participants, 45% (n=1147) reported having had a pregnancy with 38% (n=967) resulting in biological children. Participants with complex CHD were less likely to meet recommended physical activity guidelines (X^2^ (2, n = 917) = 15.9, p < 0.001), a factor that was more pronounced amongst female participants. Overall health-related quality of life was rated as good or better by 84% of participants with no difference by CHD complexity.

**Conclusion:** CHI is the largest ongoing registry of adults living with congenital heart disease in the US and includes patients with a wide variety of CHD subtypes. Many patients report mood disorders, but most report good or very good health-reported quality of life. The CHI is poised to facilitate multicenter research with the goal of improving clinical outcomes for all adults with CHD.

## Introduction

Due to tremendous advances in diagnostic and surgical techniques, the survival of patients with congenital heart disease (CHD) into adulthood has improved over the past sixty years, such that there are more adults alive with CHD than children.^1,2^ It is estimated there are over 1.5 million adults in the United States living with a congenital heart defect. The exact size of this population in the United States, and the healthcare resources needed to optimally manage their continued growth, is not well known.

Significant barriers have existed in conducting surveillance and long-term outcomes research in this patient population. The lack of a longitudinal registry in the US and the heterogeneity of CHD has resulted in an inability to fully characterize the US adult CHD (ACHD) population.^3^ In addition, there are no longitudinal registries collecting patient-reported outcomes, further contributing to the lack of knowledge in the adult CHD patient population. The lack of a robust data collection system in this population hinders research, medical advancements, and the medical community’s understanding of the population’s long-term outcomes.

To address these limitations, a multidisciplinary group of cardiac care providers and researchers, registry design experts, patient and family partners, professional society representatives, and CHD patient advocacy organizations was assembled to design the Congenital Heart Initiative (CHI), the first ACHD focused registry in the United States. Built with patient guidance, the goal was to address a critical need for adult patients with CHD by providing a deeper understanding of the CHD experience, while also offering critical infrastructure for future research and clinical care initiatives. The development of the CHI highlights how digital studies can promote innovations in research and facilitate patient-partner engagement, especially in rare disease conditions.

## Methods

### Partner Engagement and Human Centered Design

Patients with CHD, family members of patients with CHD, clinicians, and researchers were engaged throughout the entire development process, starting in March 2018 (Figure 1a). Methods for registry design were rooted in a human centered design approach with the goals of (1) discovering patient needs, (2) designing a platform with patient input, and (3) delivering a registry to specifically address patient needs.^4^

**Figure 1a and 1b:**
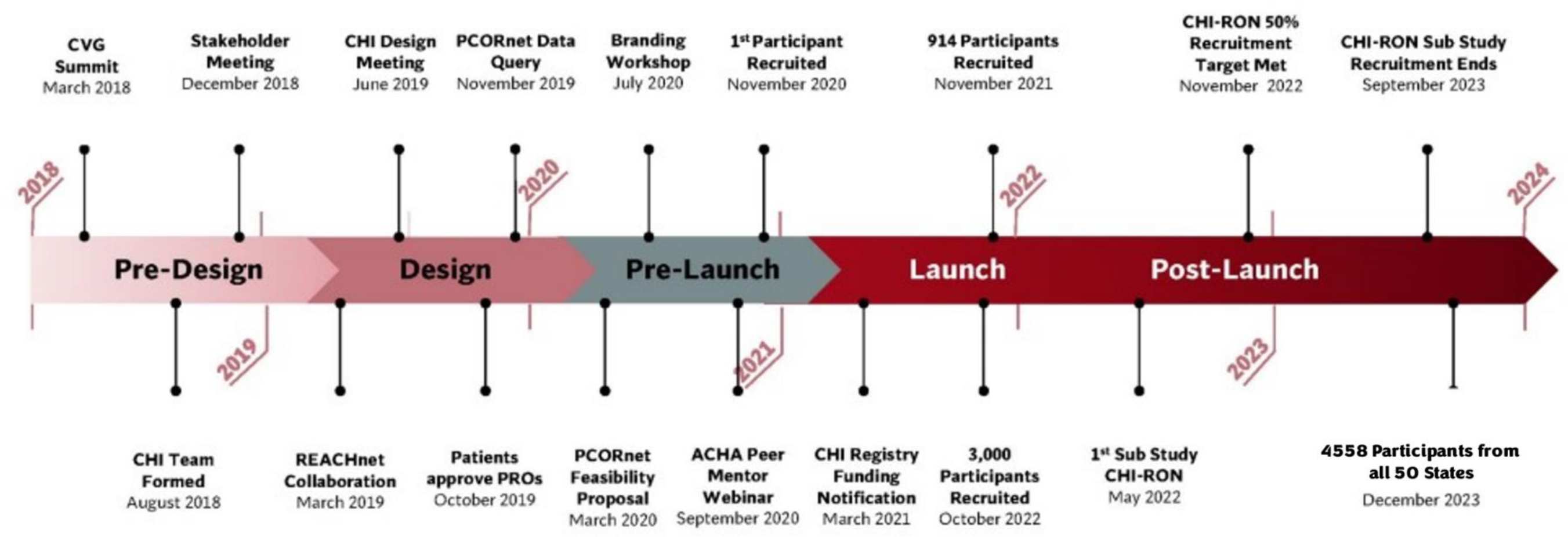
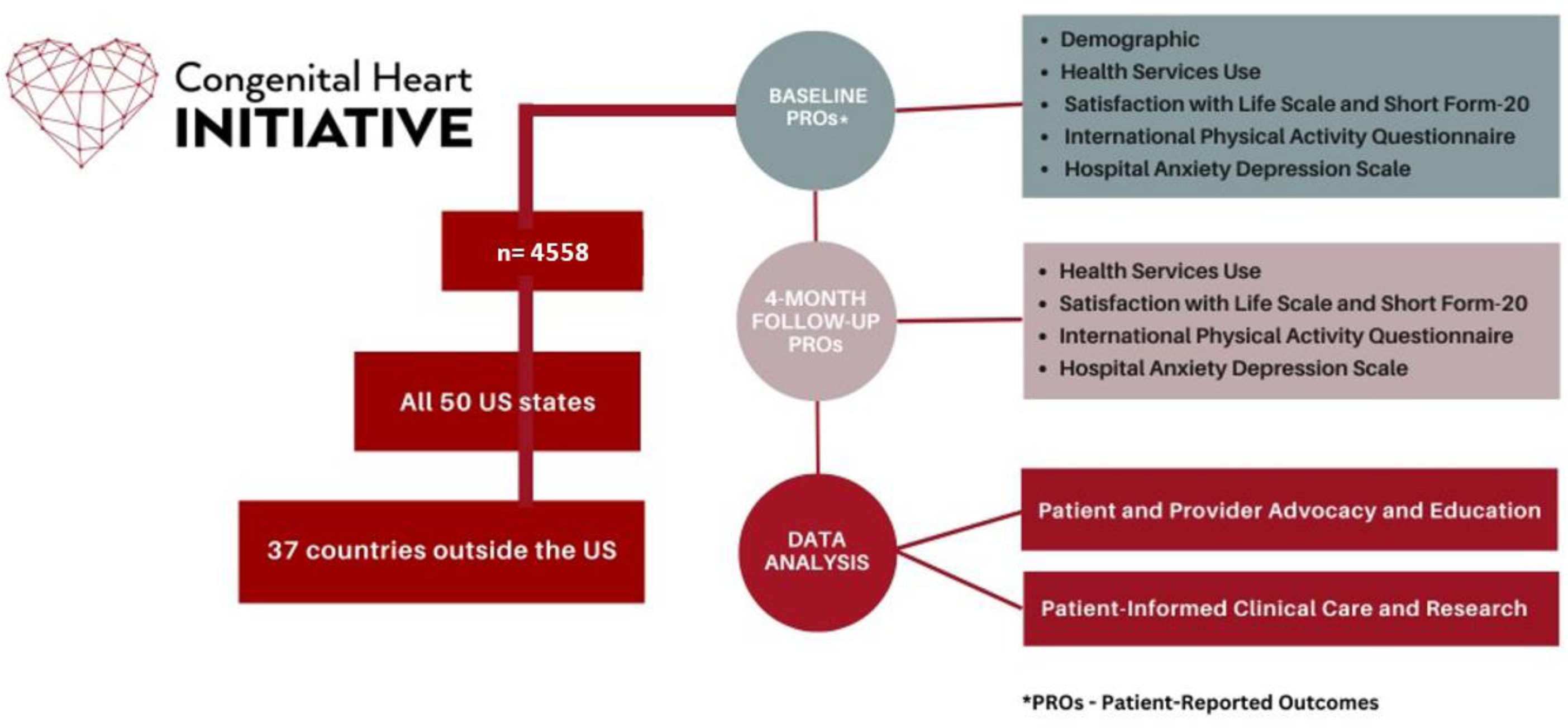
Timeline for Creation of the Congenital Heart Initiative (Figure 1a); Congenital Heart Initiative flowchart (Figure 1b).

During a patient-powered research summit (sponsored by the PCORnet Cardiovascular Health Collaborative Research Group [CRG]) that focused on participants who were adults with CHD, researchers, patients, clinicians, and representatives from the Adult Congenital Heart Association (ACHA), participants discussed research questions and outcomes that were important to CHD patients and their families. The lack of a longitudinal registry was identified as a barrier to accomplishing many of the goals prioritized by patients.

Following this summit, a group of key experts in surveillance research, registry design and management, adult congenital cardiology, quality improvement, and key patient partners and leaders from the ACHA was assembled. Additional input regarding the purpose and goals of the registry was solicited via surveys from patient and provider partners. The invitation to participate in the survey was emailed out to patients by the ACHA and through professional networking. The survey was electronically administered through Survey Monkey and all responses were anonymous. Following the synthesis of the results, an in-person meeting was held in December 2018 with sponsorship from the Adult Congenital Pediatric Cardiology section of the American College of Cardiology. Plans were established for building the registry with discussion on tool selection and testing, website design, and a branding workshop that were completed over the following year. As a result of the branding workshop (conducted by CURA strategies) the name of the registry, the Congenital Heart Initiative, was selected by the patient and provider community as a reflection of the vision, mission, and purpose of the registry.^4^

### Registry Tool Selection

Patient reported outcome (PRO) tools were initially selected by the provider and researcher community. Through an electronic survey, 13 providers/researchers narrowed down the tool selection in the domains of mental health (four tools), quality of life (three tools), and physical activity (one tool). From August through October 2019, opinions were solicited on the selected PRO tools from CHD patients and family members through web-based, electronic surveys asking for input regarding length of the survey, ability to understand the questions, and which survey best captured their information and current state. The surveys were electronically administered through REDCap and all responses were anonymous. Once the tools were narrowed down to 2-3 per domain, additional input (survey preference, time for completion, and usability) was obtained from patients through outreach from ACHA.

The final set of survey tools included a set of six baseline surveys (initial visit) and five follow-up surveys delivered every four months (Figure 1b). Baseline surveys included demographics, health services use, and four validated tools including the Quality-of-Life Short Form – 20,^5^ Satisfaction with Life Scale,^6^ mental health (Hospital Anxiety and Depressions Scale),^7^ and physical activity (International Physical Activity Questionnaire).^8^ Patients specified that e-Visits should not be longer than 10 to 15 minutes; in response, the initial intake e-Visit takes only 10 to 15 minutes to complete, while subsequent follow-up e-Visits that are delivered every four months can be completed within less than 10 minutes.

Patient reports regarding sociodemographic factors and health service use were collected through surveys that were adapted from the Congenital Heart Survey To Recognize Outcomes, Needs, and well-beinG (CH-STRONG), in addition to several customized questions designed by the study team.^9^ These questions consisted of a combination of multiple choice and multiple entry responses as well as open-ended text entry fields. In addition, COVID-19 specific questions were utilized from the COVID-Citizen Science Study, also housed on the Eureka Platform.^10^ CHD anatomy was collected using a multiple-entry check field variable with an open-ended response and then categorized (Table 1) in a hierarchical structure, along with CHD anatomy classification, as simple (I), moderate (II), or great complexity (III) as previously defined.^11^

**Table 1.** Summary of CHI Patient-Reported CHD Anatomy and Hierarchical Categorization

| **Patient-Reported Congenital Heart Defect** | **CHI Congenital Heart Disease Hierarchy** | **ACHD-AP Classification<sup>1</sup>** |
| --- | --- | --- |
| Eisenmenger syndrome/cyanotic heart disease | 1. Eisenmenger syndrome/cyanotic heart disease | III |
| Hypoplastic left heart syndrome (HLHS) | 2. Hypoplastic left heart syndrome (HLHS) | III |
| Fontan or Single ventricle or Double inlet left ventricle or Tricuspid atresia | 3. Univentricle | III |
| Transposition of great arteries (TGA) or Double outlet right ventricle (DORV) | 4. Transposition of great arteries (TGA) or Double outlet right ventricle (DORV) | III |
| Tetralogy of Fallot (TOF) or Pulmonary atresia (with VSD) and Ventricular septal defect (VSD) | 5. Tetralogy of Fallot (TOF) | TOF |
| Truncus arteriosus | 6. Truncus arteriosus | III |
| Atrial septal defect (ASD)- Pulmonary arterial hypertension (PAH) | 7. Atrial septal defect (ASD)- Pulmonary arterial hypertension (PAH) | II |
| Atrioventricular septal defect | 8. Endocardial cushion | II |
| Anomalies of Aorta | 9. Anomalies of Aorta | II |
| Coarctation of aorta (CoA) | 9. Coarctation of aorta (CoA) | II |
| Ebstein anomaly | 10. Ebstein anomaly | II |
| Pulmonary atresia (PA) (alone) or pulmonary valve stenosis (PV) | 11. Anomalies of Pulmonary atresia (PA) or pulmonary valve stenosis (PV) | II |
| Anomalies of great veins or pulmonary veins (e.g., PAPVR, TAPVR, Scimitar, etc.) | 12. Anomalies of veins | II |
| Subaortic stenosis | 13. Subaortic stenosis | II |
| Patent ductus arteriosus (PDA) | 14. Patent ductus arteriosus (PDA) | I |
| Ventricular septal defect (VSD) | 15. Ventricular septal defect (VSD) | I |
| Aortic valve stenosis or Bicuspid aortic valve | 16. Anomalies of aortic valves (AV) | I |
| Atrial septal defect (ASD) | 17. Anomalous coronary artery | I |
| Other (open-ended text entry) | 18. Other anomalies |  |
| Don't know | 19. Unknown |  |

1. Patient-reported CHD anatomy is classified as simple (I), moderate (II), or “great complexity” (III) as defined by the American College of Cardiology and the American Heart Association (Stout, 2019).

### Study Design, Setting, and Participants

The Congenital Heart Initiative (https://chi.eurekaplatform.org) is a longitudinal cohort study conducted on the Eureka Research Platform, delivered through a web-based system. The CHI was launched, after a two-year-period of intensive partner engagement, design, and testing by the core research team and patient volunteers (Figure 1). Participant enrollment is currently ongoing.

Eligibility requirements include (1) being ≥ 18 years old, (2) having a CHD diagnosis, (3) ability to participate in English, and (4) being able to provide consent and complete surveys independently. Participants are then asked to complete a baseline set of demographic surveys and four validated tools. A brief health services update survey and the four validated tools are then re-delivered every four months. The CHI was reviewed and approved by the Children’s National Hospital Institutional Review Board (IRB) Pro00016403 and built through the Eureka platform, which has been reviewed and approved by the University of California-San Francisco IRB. All participants provide electronic consent. There are no monetary incentives for participation.

### Eureka Research Platform, Data Collection and Management

The CHI is an entirely digital study hosted on the Eureka Research Platform, which has standardized elements for electronic consent, surveys, and feedback that can be customized for individual studies.^12^ Studies on the Eureka platform allow for changes in response to participant feedback, new scientific or public health findings, and new research questions. The participant can go back and change responses within a survey, but they cannot change responses after submitting the survey. Study data is stored on private, secure, Health Insurance Portability and Accountability Act (HIPAA) – compliant cloud-based servers. Data access is restricted to authorized study personnel.

### Recruitment

CHI recruitment began locally with a pre-launch period in November 2020 and was launched nationally December 7, 2020 through multiple mechanisms via collaborations with both patient and clinical partners. The CHI study was broadly advertised through webinars to the CHD patient community, press releases, and social media feeds. Toolkits were created for both organizations (i.e., clinical care centers, professional organizations, and patient advocacy organizations) and patients to further amplify the message (https://linktr.ee/CHIToolkits[linktr.ee]). Through a strong partnership with the Adult Congenital Heart Association (ACHA), webinars were created for the patient and provider community in addition to educational sessions for patient ambassadors, clinical providers, and other interested organizations. Information about the CHI, including the link to the website and instructional videos, is posted on the ACHA website. Participants were also recruited from other Eureka platform studies. In May 2021, an email was sent to 2,443 Health eHeart participants who self-identified as having CHD with information about the CHI registry. To date, there are a total of 155 CHI participants who had participated in Eureka studies prior to the launch of the CHI. In addition, targeted recruitment was performed through the Congenital Heart Initiative: Redefining Outcomes and Navigation to adult centered care (CHI-RON) study, starting May 2022. CHI-RON is the first sub-study of the CHI that involved recruitment at 12 PCORnet clinical centers.

### Registry Governance

The CHI governance is led by a multi-disciplinary advisory board of patients with CHD, clinicians, researchers, data scientists, federal funding partners, and patient advocacy organizations. The advisory board is comprised of a patient sub-committee and a scientific sub-committee. The patient sub-committee focuses on study recruitment, patient engagement, and research topic prioritization, while the scientific committee oversees project proposals and data analysis plans. As a primary goal of the registry was to facilitate additional research, an intake process was developed for data requests from the scientific community requiring advisory board and executive committee approval prior to the release of data.

### Statistical Analysis

Data processing, harmonization, and analysis were conducted by an accredited data science team data science team at Children’s National Hospital. Categorical variables collected at baseline and follow-up e-Visits are presented as absolute numbers and percentages and include sociodemographic, clinical, and behavioral variables collected from each participant including, but not limited to, age, sex, personal and household income, race and ethnicity, CHD anatomy, education level, cardiac device implantation, history of surgery, physical and mental comorbid conditions. Categorical variables were presented as frequencies and percentages to provide a comprehensive overview. Descriptive statistics were employed to understand the associations between various factors, including the complexity of heart defects (classified as simple, moderate, or complex), physical activity levels, mental health comorbidities, and socioeconomic and healthcare access variables. All categorical variables were analyzed using Chi-square or Fischer’s exact test as appropriate. All the assumptions were assessed. All analyses were done using SAS 9.4 (SAS Institute., Cary, NC, USA).

## Results

### Registry Tool Selection

PRO tools were initially selected based on patient goals for the registry as previously described.^4^ The final PRO tools were selected by a total of 177 (69.5% female) CHD patients, family members, and guardians. Most respondents (70.1%) were recruited to take part in the survey by the ACHA and identified as CHD patients (74%) with an average age of 46 years +/- 14.8. A total of 55 individuals participated in the survey regarding the utility of the mental health tools PHQ-4, PHQ-8, GAD-7, and HADS for measuring PROs relating to anxiety and depression ^7,13,14^. Overall, the highest proportion of respondents indicated that the HADS best measured their anxiety and depression (41%). A total of 122 participated in the survey regarding the utility of the SF-36, PROMIS-29, and the IAQ tools for measuring their quality of life^5,15,16^. Overall, the highest proportion of respondents indicated that the SF-36 best measured their quality of life (46.7%). The IPAQ was the only physical activity PRO tool assessed.

### Registry Demographic Data

As of December 31, 2023, a total of 4558 participants have enrolled in the Congenital Heart Initiative (Table 2); 88% of participants completed their initial visit and 62% of participants have completed at least one follow-up visit. There were a higher percentage of female participants (2538, 56%) with an average age of the total participants in the cohort of 38.5 +/-13.9 years. The participant distribution by sex and patient-reported CHD is shown in Figure 2.

**Figure 2.**
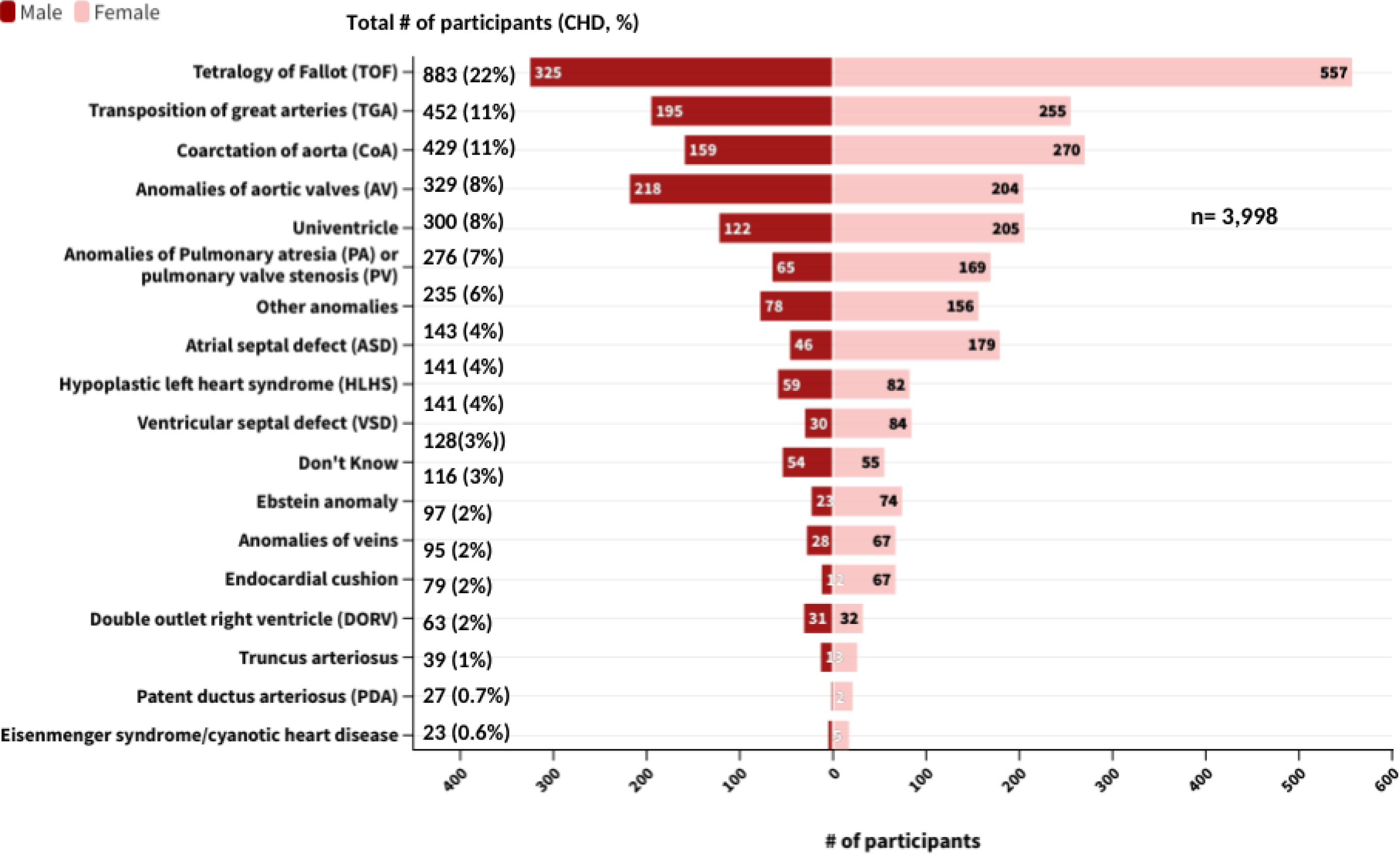
Congenital Heart Initiative participants who joined by December 31, 2023, by sex assigned at birth and patient-reported congenital heart disease anomaly (n = 3998, completed initial visit).

**Table 2.** Summary of Participant Characteristics enrolled through December 31, 2023

|  |  | **All, *n*** | **Male, *n* (%)** | **Female, *n* (%)** | **Other, *n* (%)** | **Unknown, *n* (%)** |
| --- | --- | --- | --- | --- | --- | --- |
| **Sex at birth** |  | 4558 (100%) | 1466 (32%) | 2530 (56%) | 2 (0.04%) | 560 (12%) |
| **Age** |  |  |  |  |  |  |
|  | Median |  |  |  |  | - |
|  | 18-25 | 732 (16%) | 287 (6%) | 444 (10%) | 1 (0.02%) | - |
|  | 26-35 | 1180 (26%) | 434 (10%) | 746 (16%) | - | - |
|  | 36-45 | 991 (22%) | 346 (8%) | 645 (14%) | - | - |
|  | 46-55 | 532 (12%) | 169 (4%) | 362 (8%) | 1 (0.02%) | - |
|  | 56-65 | 341 (7%) | 136 (3%) | 205 (4%) | - | - |
|  | over 65 | 207 (5%) | 89 (2%) | 118 (3%) | - | - |
|  | Unknown | 575 (12%) | 5 (0.1%) | 10 (0.2%) | - | 560 (12%) |
| **Marital Status** |  |  |  |  |  |  |
|  | Single | 1796 (39%) | 698 (15%) | 1097 (24%) | 1 (0.02%) | - |
|  | Divorced | 224 (5%) | 59 (1%) | 165 (4%) | - | - |
|  | Married | 1861 (41%) | 678 (15%) | 1182 (26%) | 1 (0.02%) | - |
|  | Other | 116 (3%) | 31 (1%) | 85 (2%) | - | - |
|  | Unknown | 561 (12%) | - | 1 (0.02%) | - | 560 (12%) |
| **Children** |  |  |  |  |  |  |
|  | Yes, biological | 1517 (33%) | 549 (12%) | 967 (21%) | 1 (0.02%) | - |
|  | Yes, adopted | 94 (2%) | 20 (0.4%) | 74 (1.6%) | - | - |
|  | No | 2306 (51%) | 874 (19.2%) | 1431 (31.4%) | 1 (0.02%) | - |
|  | Other | 81 (2%) | 23 (1%) | 58 (1%) | - | - |
|  | Unknown | 560 (12%) | - | - | - | 560 (12%) |
| **Race and ethnicity** |  |  |  |  |  |  |
| **Hispanic** |  | **386 (8.5%)** | **128 (2.9%)** | **257 (5.6%)** | **1 (0.02%)** | - |
|  | Cuban | 51 (1.2%) | 16 (0.4%) | 35 (0.8%) | - | - |
|  | Mexican | 92 (2%) | 27 (0.6%) | 65 (1.4%) | - | - |
|  | Puerto Rican | 77 (1.7%) | 27 (0.6%) | 50 (1.1%) | - | - |
|  | Other Hispanic | 166 (3.6%) | 58 (1.27%) | 107 (2.35%) | 1 (0.02%) | - |
| **Non-Hispanic** |  | **3590 (78.8%)** | **1330 (29.2%)** | **2259 (49.6%)** | **1 (0.02%)** | - |
|  | Asian | 138 (3%) | 65 (1.4%) | 73 (1.6%) | - | - |
|  | Native American | 10 (0.21%) | 3 (0.06%) | 7 (0.15%) | - | - |
|  | Multi-race | 265 (6%) | 84 (2%) | 181 (4%) | - | - |
|  | Pacific Islander | 1 (0.02%) | - | 1 (0.02%) | - | - |
|  | White | 3139 (69%) | 1166 (26%) | 1972 (43%) | 1 (0.02%) | - |
|  | Don't Know | 6 (0.14%) | 3 (0.07%) | 3 (0.07%) | - | - |
|  | Other | 26 (0.6%) | 9 (0.2%) | 17 (0.4%) | - | - |
|  | White, Unknown |  |  |  |  |  |
|  | Hispanic | 5 (0.1%) | - | 5 (0.1%) | - | - |
| **Unknown** |  | **582 (12.5%)** | **8 (0.18%)** | **14 (0.31%)** | - | 560 (12%) |

**Table 2 (continued). Summary of Participant Characteristics enrolled through December 31, 2023**
|  | **All, *n* (%)** | **Male, *n* (%)** | **Female, *n* (%)** | **Other, *n* (%)** | **Unknown, *n* (%)** |
| --- | --- | --- | --- | --- | --- |
| **Education** |  |  |  |  |  |
| Never attended school or only attended kindergarten | 1 (0.02%) | 1 (0.02%) | - | - | - |
| 9<sup>th</sup> to 12<sup>th</sup> grade | 74 (1.6%) | 31 (0.7%) | 43 (0.9%) | - | - |
| High school graduate, GED, or alternative | 427 (9.4%) | 173 (3.8%) | 253 (5.6%) | 1 (0.02%) | - |
| Some college, no degree | 703 (15%) | 253 (5%) | 450 (10%) | - | - |
| Associate degree | 308 (7%) | 89 (2%) | 219 (5%) | - | - |
| Bachelor's degree | 1264 (28%) | 477 (11%) | 787 (17%) | - | - |
| Graduate or professional degree | 1217 (27%) | 441 (10%) | 775 (17%) | 1 (0.02%) | - |
| Don't know/ not sure | 4 (0.1%) | 1 (0.03%) | 3 (0.07%) | - | - |
| Unknown | 560 (12%) | - | - | - | 560 (12%) |
| **Personal Income** |  |  |  |  |  |
| \$0 - \$24,999 | 780 (17%) | 282 (6%) | 497 (11%) | 1 (0.02%) | - |
| \$25,000 - \$49,999 | 454 (10%) | 150 (3%) | 304 (7%) | - | - |
| \$50,000 - \$99,999 | 738 (16%) | 300 (6%) | 438 (10%) | - | - |
| \$100,000 - \$149,999 | 273 (6%) | 130 (3%) | 143 (3%) | - | - |
| \$150,000 and greater | 248 (5.4%) | 165 (3.6%) | 82 (1.8%) | 1 (0.02%) | - |
| Prefer not to answer | 291 (6.3%) | 111 (2.4%) | 180 (3.9%) | - | - |
| I don't know | 107 (2.3%) | 26 (0.5%) | 81 (1.8%) | - | - |
| Unknown | 1667 (37%) | 302 (7%) | 805 (18%) | - | 560 (12%) |
| **Household Income** |  |  |  |  |  |
| \$0 - \$24,999 | 231 (5%) | 90 (2%) | 141 (3%) | - | - |
| \$25,000 - \$49,999 | 236 (5.1%) | 93 (2%) | 143 (3.1%) | - | - |
| \$50,000 - \$99,999 | 608 (13%) | 224 (5%) | 383 (8%) | 1 (0.02%) | - |
| \$100,000 - \$149,999 | 431 (9.4%) | 166 (3.6%) | 265 (5.8%) | - | - |
| \$150,000 and greater | 711 (15.6%) | 336 (7.4%) | 374 (8.2%) | 1 (0.02%) | - |
| Prefer not to answer | 340 (7.5%) | 134 (3%) | 206 (4.5%) | - | - |
| I don't know | 334 (7.4%) | 121 (2.7%) | 213 (4.7%) | - | - |
| Unknown | 1667 (37%) | 302 (7%) | 805 (18%) | - | 560 (12%) |

Participants enrolled in the study represented all 50 states with the highest proportion of patients from New York (463, 14%), Florida (393, 12%), and Ohio (303, 9%). A total of 1,183 participants reported their home state in the northeast (35%), 929 from southern states (28%), 594 from the western states in the US (18%), and 496 from midwestern states (15%).

Approximately 41% of participants are married and 37% had children (see Table 3). Among the participants identifying as female, 45% (n=1147) reported having a pregnancy, with 32%(n=799) of those patients reporting more than one pregnancy. The most common diagnoses of women undergoing pregnancy included: tetralogy of Fallot (24.6%), shunt lesions (13%), and coarctation of the aorta (11%), reflective of the population of CHI participants.

### Patient-Reported Outcomes Data

Of the participants followed through December 31, 2023, approximately 18% reported no co-morbidities (Figure 3). Arrhythmias were the most common cardiac co-morbidity (1310, 29%) followed by hypertension (763, 17%) and congestive heart failure (592, 13%). Mood disorders like anxiety or depression (1339, 29%), asthma (577, 13%), and anemia (467, 10%) were the most commonly reported non-cardiac complications. Thirteen percent of patients reported other chronic health problems. The top three most prevalent “other” chronic health conditions included thyroid issues, migraines, and gastric reflux issues. Most patients had undergone at least one cardiac surgery, with only 14% reporting no prior surgical intervention. Seventeen percent of participants reported having had a device such as a pacemaker or a defibrillator implanted; 6% reported having “other” devices which included stents, transcatheter prosthetic valves, and implantable recorders.

**Figure 3.**
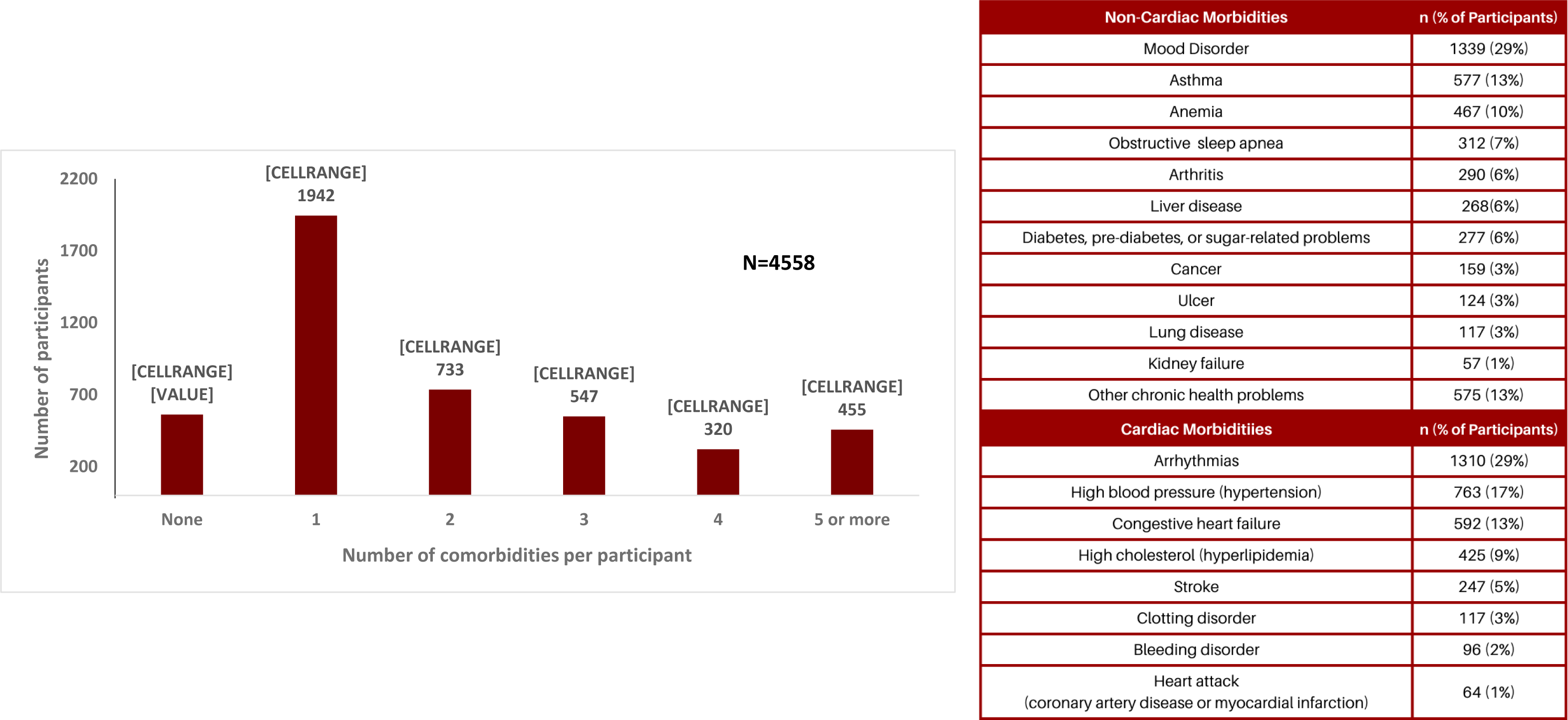
Summary of Congenital Heart Initiative patient-reported comorbidities per participant and overall cardiac and non-cardiac comorbidity prevalence (n= 4558).

Most patients reported being currently in care, with 84% (n=3,354) reporting a visit to their cardiologist within the past 2 years and 71% (n=2,841) report visiting a heart doctor in the prior year. Thirty one percent (n=1,256) of patients reported a 3+ year gap in cardiac care with the top five reasons being that they felt well and didn’t think they needed to go to the doctor (54%, n=675), moved (16%, n=198), found it too far to drive (11.2%,n=141), their parents stopped taking them (13%, n=162), or they had changed or lost insurance (14.5%, n=183). Fewer patients reported a recent primary care physician (PCP) visit with 63% (n=2,548) reporting visiting their PCP within one year. When asked about reasons for historical gaps in primary care, the top three reasons cited were that they felt well (152, 36%), they didn’t think they needed to see a PCP (27.6%, n=116%), or were moving to a different city (19.3%, n=81).

To assess whether the severity of CHD impacts exercise levels, participants were stratified by CHD complexity.^11^ Weekly physical activity intervals based on PROs were compared with recommended physical activity guidelines of 75 minutes of vigorous activity per week or a combination of vigorous and moderate activity totaling 150 minutes per week.^17^ Of the 3,321 participants with physical activity data, nearly 28% (917) met physical activity guidelines. Initial analysis of baseline data showed that females were less likely than males to meet recommended physical activity guidelines (Figure 4a). Chi-square tests of independence were performed which showed a significant association between CHD severity and the reported level of physical activity (X^2^ (2, n = 917) = 15.9, p < 0.001. When stratified by CHD anatomy, patients with complex CHD reported lower rates of meeting physical activity recommendations (25%, n = 408, p = 0.005) compared to those with moderate complexity (32%, n = 254, p < 0.001). Most patients reported a good or better health-related quality of life, regardless of CHD complexity (Figure 4b).

**Figure 4a and 4b.**
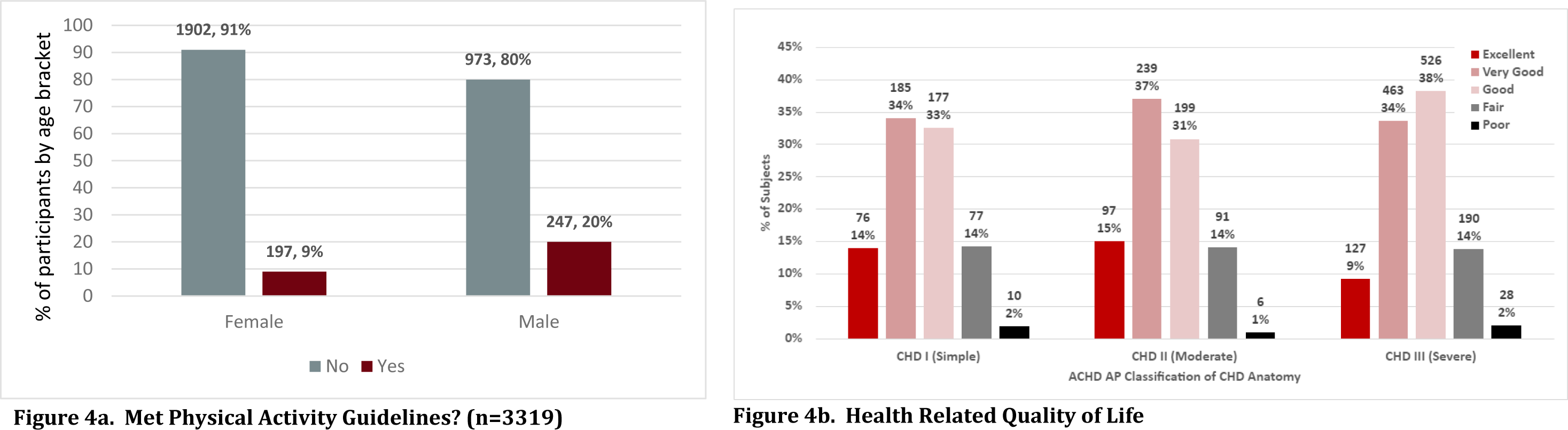
Participant physical activity by sex (Figure 4a, left) and health related quality of life by CHD complexity (Figure 4b, right). Physical activity goals are considered met if the participant achieves 75 minutes of vigorous activity per week or a combination of vigorous and moderate activity totaling 150 minutes per week (Piercy et al., 2018).

## Discussion

The overarching goal of the CHI is to improve the quality of life for all adults with CHD while also filling a previously identified gap to advance research, the lack of a longitudinal registry.^3^ The two major aims of the CHI are to: (1) improve clinical care for ACHD patients, and (2) facilitate multi-center research. The CHI seeks to improve our understanding of the CHD population, inform researchers of the needs within the community, and spur innovation with new research on treatment options. We were able to collect health data from patients from across the United States, with targeted outreach at sites participating in the CHI-RON study. Additionally, our ability to successfully recruit more than 4,500 participants over the first three years of enrollment demonstrates the feasibility and acceptability of individuals and communities to engage in this digital platform.

### Patient and Partner Engagement

One of the unique aspects of the CHI registry is the strong community engagement. The registry was developed through countless hours of volunteer effort from patients, providers, and researchers, resulting in a platform designed by the community it was intended to serve. Several strategies based on human centered design methods have been employed to promote sustained engagement and participation through sequential surveys which are often a challenge in digital clinical studies. Human centered design (HCD) is a problem-solving qualitative research framework that is grounded in empathy and understanding.^18^ We utilized this methodology to discover the needs of the community, design a registry that was customized to patients, and deliver a platform that addressed patient priorities.^4^

Additional engagement strategies are designed to meet participants’ needs. Many participants voiced a feeling of “being alone” with a desire to connect to other patients and the CHD community. One participant stated that “they had always felt forgotten, like a lost child” until they discovered the CHI. To promote sustained engagement, newsletters are sent to CHI participants addressing the need of improved connections and facilitating knowledge about CHD. The newsletters provide information about their local community events and walks, as well as recorded webinars and updates about the registry. The close partnership with the ACHA facilitates seamless distribution of information regarding these events. Connecting to the CHI starting October 2022, a virtual ACHA café is a novel technique to promote engagement within the CHD community. Facilitated by trained patient ambassadors and peer mentors from the ACHA, a virtual café was held monthly till December 2023, allowing patients to connect with each other on a virtual format through a secure videoconferencing link.

### Data Governance and Return of Results

Ethical themes and the principles of sound governance for use of Big Data were employed to develop the governance structure for the CHI framework.^19–21^ As patients consider participation in the CHI, it is important to maintain respect for the individual in the context of seeking answers that apply to and can advance care for all.^22^ As the data that comprises the CHI is provided by patients, the CHI team understands it is critical to provide the results back to the patient community. Our first and second annual reports can be accessed on the ACHA website (https://www.achaheart.org/media/4024/chiannualreport2023.pdf). To further promote ongoing engagement, data summaries releasing information to the community are included in engagement newsletters and through the ACHA website updates on a periodic basis.

Future research utilizing the data from the CHI registry and the platform of the CHI itself is one of the major goals identified by patients, clinical providers, and researchers. The CHI governance structure is guided by several committees and cores, including a steering committee and a multi-disciplinary advisory board. Importantly, patients are part of the governance board and part of the executive leadership of the registry. Currently, data summaries from the registries are reviewed by each group and are not released for distribution until both the patient and clinical providers review the format and offer clarifications. Both groups are comprised of numerous partners, including physicians, researchers, and patients. Data requests can be made through a data intake form and are then reviewed by the multidisciplinary advisory board to ensure requests are compliant with federal regulations. The advisory board then determines the next steps, although the process continues to be streamlined.

### Research Utilization and Multicenter Studies

One of the major goals of the CHI is to facilitate multicenter research, which plays a pivotal role in advancing understanding of long-term outcomes in ACHD. Increasing sample size, recruiting diverse patient populations, improving reproducibility, and facilitating clinical trials are all key advantages that multicenter research offers in the study of rare disease. The CHI provides a research ready cohort of patients while also providing key updates on the needs of ACHD patients within the United States. There are two current multicenter, sub-studies of the CHI registry, addressing the impact of gaps in recommended care for ACHD patients and establishing how to include ACHD patients with neurodevelopmental disabilities in patient centered outcomes research.

The first sub-study is called Congenital Heart Initiative: Redefining Outcomes and Navigation to adult-centered care (CHI-RON). While the CHI established a platform to collect patient-generated data, there was a lack of data integration from the clinical provider or electronic medical record. To address this need, a collaboration was forged with PCORnet (the National Patient-Centered Clinical Research Network), an integrated network with healthcare encounter data from more than 30 million people across the United States.^23^ The CHI-RON study is a first-of-its-kind registry for ACHD patients that collects real-world data (RWD) from multiple sources. To date, there are approximately 125,000 patients in the PCORnet observational cohort with only electronic medical record (EMR) data, of which, approximately 5,000 have linked claims-based data. Approximately 3,027 participants have been recruited to the CHI-RON study and the CHI. This subset of participants receiving care from 12 participating PCORnet centers will have three distinct types of data included in their profiles: patient-reported outcomes, claims, and electronic health records (EHRs). The CHI-RON sub-study seeks to understand the impact of receiving American Heart Association/American College of Cardiology guideline-directed cardiac care on long-term outcomes in adults with CHD.^11^

A second sub-study, Achieving Equity: Inclusion of Adults with Congenital Heart Disease Living with Neurodevelopmental Disability (NDD) in Patient Centered Outcomes Research aims to improve access for individuals with neurodevelopmental disabilities and CHD. Unfortunately, a key demographic that remains underrepresented in the CHI is patients with NDD. To be eligible for the CHI, participants must be able to complete the PRO tools offered through the CHI independently. As a result, individuals with CHD and NDD are often ineligible to participate, representing a major gap in PRO research, as nearly a third of the population are excluded. To address this gap, we plan to modify our existing successful CHI engagement rubric (based on the PCORI engagement rubric) to create an alternate pathway for inclusion of patients with CHD and NDD to participate in the CHI.

Additional studies are in development focusing on three key topics identified by the larger CHD community as priority areas for the CHI to explore. These include the healthcare needs to reduce maternal morbidity in individuals with CHD, virtual interventions for additional mental health support, and the evaluation of treatment modalities for CHD patients.

### Limitations

One of the major limitations of the registry is that it currently contains PRO data only. Recall bias, underlying neurocognitive challenges, and survey fatigue can all limit continued participation in the CHI. There is also participant bias as individuals concerned about their health issues and those who have access to and are more comfortable with technology are likely overrepresented. There is a higher percentage of female participants, with larger distributions in certain parts of the country. In addition, there is a lack of diversity with most participants identifying as non-Hispanic white.

To address these issues, a major goal of the CHI is to increase racial and ethnic diversity and geographic distribution of participants. The CHI-RON sub-study seeks to address these issues by generating recruitment lists that prioritize the out-of-care population in addition to under-represented minorities. Further engagement strategies are being developed to advertise the study across the country to a more diverse population.

### Future Directions

Future directions of the CHI include improving engagement, including partnerships with other CHD-focused registry platforms that are currently lacking in PRO metrics. These partnerships will be needed to develop a sustainability plan, allowing the registry to continue to flourish. Additional goals include increasing the participation of diverse populations while promoting the use of the CHI for education, data dissemination, and patient-centered research. The CHI is moving toward these goals through (1) increasing social media content to solicit research priorities from the patient community; (2) leveraging existing partnerships with the ACHA for improved data dissemination to the patient and provider community through web-based and video content; and (3) working with interested researchers to harness the existing data within the CHI while also developing new projects. Our hope is that researchers will use the CHI as a recruitment tool for their studies. In addition, with the CHI-RON sub-study, future models for simulated clinical trials and risk prediction scales can hopefully be developed for this population of patients in whom clinical trials have been historically difficult to conduct. Finally, the goal to make this a lifelong registry extending into the pediatric population and the neurodivergent population remain but will require additional engagement and infrastructure development.

### Conclusion

In summary, the CHI is an essential tool and the first step for helping to deliver improved outcomes for all ACHD patients. Designed and fueled by patients, the CHI registry serves as a tool to collect PROs while also soliciting the needs of the patient community regarding future research. It also demonstrates the feasibility of the CHI registry as a tool that unites the community of ACHD patients, providers, and researchers. CHI participants continue providing valuable insight into aspects of their daily lives as ACHD patients. As the CHI continues to grow and develop, increasing the diversity of participants will be critical to understanding the needs of the ACHD community across the United States. Additional partnerships with other organizations, continued innovation in data usage, and improved engagement will continue to spur innovation and future research aimed at the improved quality of care for all ACHD patients.

## Data Availability

Data is embargoed pending publication. For further inquiries, please

## Abbreviations

CHI: Congenital Heart Initiative
CHD: Congenital Heart Disease
HIPAA: Health Insurance Portability and Accountability Act
PROs: Patient Reported Outcomes
ACHA: Adult Congenital Heart Association
CHI-RON: Congenital Heart Initiative: Redefining Outcomes and Navigation to adult centered care
PCORnet: National Patient-Centered Clinical Research Network
EMR: Electronic Medical Records
HCD: Human centered design
NDD: Neurodevelopmental Disability

## Special acknowledgement

The authors would like to acknowledge the incredible contributions of Carol Maguire, RN; her passion and dedication to the ACHD community was a driving force for the creation of a registry exclusively for ACHD patients. In addition, we would like to acknowledge the following individuals: Danielle Hile, Susan Timmins, Alison Gill, Ryan O’Connor, Ken Woodhouse, John Latsha, Lena Morsch, Kristin Downing, Elisa Bradley, Ari Cedars, Abigail Khan, Karen Uzark, and the countless patients, researchers and clinical providers who provided valuable input and time to create and grow the CHI.

